# Dynamic Individual Prediction of Conversion from Mild Cognitive Impairment to Probable Alzheimer’s Disease using Joint Modeling

**DOI:** 10.1101/2024.07.15.24310224

**Authors:** Abderazzak Mouiha, Olivier Potvin, Simon Duchesne, the Alzheimer’s Disease Neuroimaging Initiative

## Abstract

**Background:** We propose a joint model predicting the risk of conversion from MCI to AD that considers the association between biomarker evolution and disease progression.

**Methods:** We selected 814 MCI subjects (285 progressives, 529 stables) who had at least four follow-up MRI visits from the ADNI dataset. The values of Alzheimer’s Disease Assessment Scale-Cognitive (ADAS-Cog) were used as a surrogate of time. A mixed linear model was fitted for bilateral hippocampal volumes (HC) versus ADAS-Cog, education, age and sex and a Cox model for risk progression. The association between HC evolution and risk conversion was estimated by fitting a joint model.

**Results:** Our results show (1) significant association (*p* < .0001, C.I.= [0.0864; 0.1217]) between bilateral HC and risk of conversion; (2) on average, the risk of progression increased as HC decreased; and (3) the individual prediction of the risk is dynamic, i.e., updated at each follow-up. The AUC of our model for the whole group increased to reach 0.789 at the last follow-up.

**Conclusions:** Applicable to AD and generalizable to other biomarkers and covariates, this joint methodology has a direct application in the clinical estimation of individual risk.

## 1. INTRODUCTION

Dementias, for which Alzheimer’s disease (**AD**) is the primary aetiology, are at the forefront of elderly’s health concerns (1), with evidence showing that risk doubles every five years after 65 years of age (2). In fact, sufficient evidence has been gathered from biomarker studies as well as post-mortem, pathological reports to postulate that the course of pseudo-sporadic AD spans more than two decades (3), echoing evidence collected in studies of autosomal-dominant AD (4), with a multiplicity of risk factors involved (5), including sex (6).

As one moves earlier in disease stages from the point at which dementia can be diagnosed, accurate prognosis becomes increasingly problematic. A large body of literature has focused on extracting biomarkers of interest from genetics, neuroimaging, and fluids, that have identified a prodromal clinical phase for AD - mild cognitive impairment (**MCI**). Models have been developed demonstrating and predicting the high risk of developing dementia for this group (7–9). However, such models, now overwhelmingly relying on machine learning, while providing an estimate of the group-wise risk of progression, tend to fare less well at individual prognosis and prediction of individual outcomes. Hence, this information is but of little value clinically, where physicians confront individual patients. Therefore, having a model that could dynamically predict an individual’s probability of progression to AD, based on a priori information, would be valuable.

Statistical methods for separate analysis of outcomes (e.g., conversion to AD, death) are well established in the literature (e.g., (10)), especially when the association between longitudinal and time-to-event (in our case, conversion from MCI to AD) is significant. It should be noted however that an extended Cox model is not appropriate when the time-dependent covariate is internal (i.e., endogenous covariate) and changes values between follow-up visits. Indeed, it is not reasonable to assume that biomarkers remain constant between follow-ups [(11), section 3.5]. Joint statistical models account for this special feature of endogenous covariates and the association between time-to-event conversion to treat the information simultaneously, and therefore provide valid and efficient inferences (12).

Our objective is to present such a joint longitudinal-survival model (13) to assess the association between longitudinal biomarkers and the event of interest (i.e., progression to clinically probable AD), taking into account the special feature of the endogenous covariate. The outcome of the model will be a dynamic prediction of time of conversion for a new subject and an “average” prediction for a cohort.

To demonstrate the applicability of our model, we focused on one of the most studied biomarkers in AD, namely hippocampal (**HC**) volume. Antemortem HC volumes measured on magnetic resonance imaging (**MRI**), especially bilateral HC volumes, have been correlated with dementia severity and density of hippocampal neurofibrillary tangles at autopsy (14). In MCI converting to probable AD, progressive HC atrophy is predictive of conversion (18). To build our model and test our hypotheses, we relied on data from the Alzheimer’s disease Neuroimaging Initiative (**ADNI**).

## 2. METHODS

### 2.1 Study design and participants

Data used in the preparation of this article were obtained from the Alzheimer’s Disease Neuroimaging Initiative (ADNI) database (adni.loni.usc.edu). The ADNI was launched in 2003 as a public-private partnership, led by Principal Investigator Michael W. Weiner, MD. The primary goal of ADNI has been to test whether serial magnetic resonance imaging (MRI), positron emission tomography (PET), other biological markers, and clinical and neuropsychological assessment can be combined to measure the progression of mild cognitive impairment (MCI) and early Alzheimer’s disease (AD). For up-to-date information, see www.adni-info.org The ADNI study has been collecting time-to-event data, including the progression from MCI to probable AD, as well as the conversion of cognitively healthy subjects (CTRL) to MCI or probable AD. Additionally, it has been collecting repeated longitudinal measures, at intervals of 6-12 months on a host of biomarkers for each patient, such as HC volumes, but also other indicators of neurodegeneration.

We downloaded the data used in the preparation of this article from the ADNI database on July 13^th^, 2022. We concentrated our analysis on the 814 individuals categorized as MCI in either ADNI phases (ADNI1, ADNI2/GO, ADNI3) who had been followed for at least four years.

Participants with MCI were divided into stable (MCIs) and progressing (MCIp) groups according to the latest clinical diagnostic available within follow-up window.

### 2.2. Measures and variables

At each visit, clinical diagnosis was assessed according to National Institute of Neurological and Communicative Disorders and Stroke and the Alzheimer’s Disease and Related Disorders Association criteria for probable AD (19). Individuals were also evaluated using the Alzheimer’s Disease Assessment Scale-Cognitive 13-item version (**ADAS-Cog** (20); range, 0-84 points), which allows to adequately track individuals from normal cognitive status to MCI and AD (21). High ADAS-Cog scores indicate of poorer cognitive impairment.

Predictors in joint models must be decreasing in value, and so ADAS-Cog scores were flipped. Further, ADAS-Cog were used as a time equivalent variable given their relevance at tracking cognitive status through the disease trajectory.

We used bilateral HC volumes generated by FreeSurfer 6.0 (https://surfer.nmr.mgh.harvard.edu/). Following FreeSurfer processing, brain segmentations were visually inspected through at least 20 evenly distributed coronal sections, then HC volumes were adjusted for image quality and intra-cranial volumes through NOMIS (22). Briefly, NOMIS adjust the segmentation volumes according to estimated intra-cranial volume, contrast-to-noise ratio of the image, and the total number of defect holes over the whole cortex, i.e. topological errors in the initial cortical surface reconstructions, through regression models based on nearly 7000 cognitively healthy adults. The NOMIS Z scores were obtained by subtracting the NOMIS model’s predicted value from the true value divided by the root mean square error of the NOMIS model. These Z scores were then used in our joint model.

To fulfill the requirements of our model, we selected only subjects that were not missing ADAS-Cog or MRIs at each timepoint.

### 2.3 Statistical analysis and joint model structure

Our research question was “how can evolution in HC volumes (longitudinal measurements) as an internal (endogenous) covariate predict the risk of progression from MCI to AD?”. The primary outcome in this study became therefore conversion from MCI to AD, and the longitudinal measurements were HC volumes. To answer our research question, the association between HC volume evolution and progression from MCI to AD was estimated (Table 2).

### 2.4 Joint model formulation

The extended Cox model is only appropriate for exogenous time-dependent covariates (e.g., age, air pollution, etc.) and therefore cannot handle endogenous longitudinal biomarkers (e.g., HC volumes) (11). When the primary interest is the association between such biomarker and conversion to AD, a joint model can account for the special feature of this endogenous covariate. Based on linear mixed-effects and relative risk conversion models, we constructed the joint model in two parts (23):

#### (1) Part one - Linear mixed-effects model

The longitudinal change in HC volumes can be assessed using the following linear mixed model:

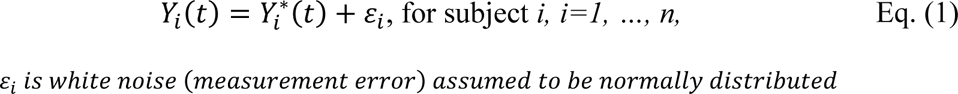

where *Y*_*i*_(*t*) is the observed longitudinal bilateral HC measure for subject *i* at time point *t*, and 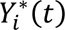, the unobserved (true underlying) value of bilateral HC, that is obtained by the following equation:

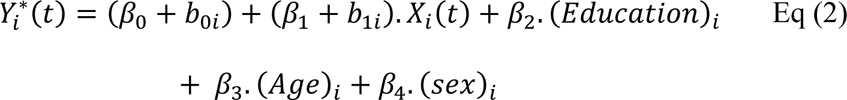

where *β*_0_, *β*_1_, *β*_2_, *β*_3_, *β*_4_ are fixed effects, *b*_0*i*_, *b*_1*i*_ are random effects (intercept and slope), *X*_*i*_(*t*) represents the biomarker trajectory for subject *i* at time *t*. *Education*, *Age* and *Sex* were used as covariates.

#### (2) Part two - Survival model

The risk of conversion from MCI to AD can be assessed by the following Cox model:

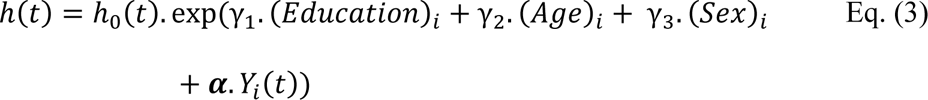

where *h*(*t*) measures the risk of conversion as a function of the longitudinal biomarker, education, age and sex; *h*_0_(*t*) is the common risk to all subjects included in the study; *γ*_*i*_, *i* = 1, 2, 3. are the effect of W=(*Education*, *Age, Sex),* as indicated in Figure 1, on conversion time; and ***α*** measures the association between longitudinal biomarker values and time-to-conversion. A value of ***α*** = 0 means that there is no association between longitudinal biomarker values and the risk for conversion, so the information from biomarker changes does not improve the estimation of the progression probability, and one uses only the analysis based on the time-to-conversion. We used a *t*-test to verify the association between HC volume changes and time-to-conversion and measured the strength of association by a Wald test.

**Figure 1.**
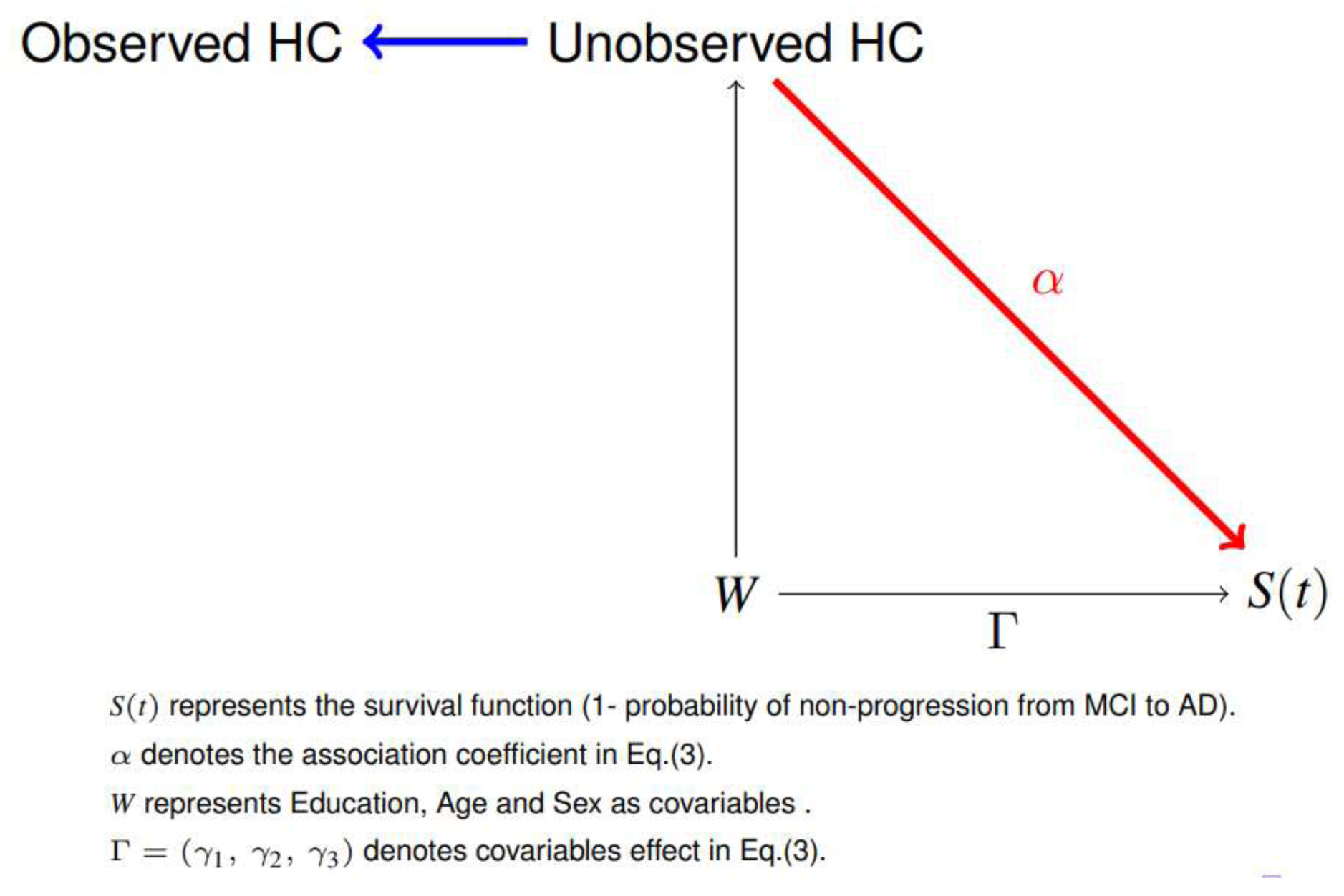
Causal diagram for the joint model. If *⍺* = 0, the analysis using joint modeling is equivalent to separate analyses.

#### (3) Longitudinal and survival combined models: joint modeling

From equations (1) to (3) above, and (24) the density of joint model is given by:

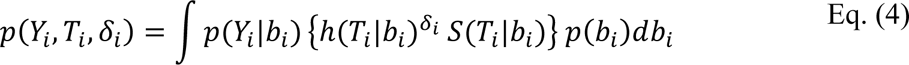

where *T*_*i*_ is the observed time-to-conversion for patient *i; δ*_*i*_ is the event indicator equals 1 if a subject *i* progressed to AD and 0 if not; 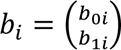 is a vector of random effects that explains the interdependencies; p(.) density function and S(.) survival function. The joint model in eq. (4) can be represented by the causal diagram shown in Figure 1 (25

#### (4) Dynamic prediction

We focused on the conditional probability of not converting to AD at time *u* ≥ *t* , given that the subject *j* was also MCI at time *t*, *t* ≥ 0. This probability is conditional on available measurements of bilateral HC up to time *t:* ℳ_*j*_(*t*) = {*Y*_*j*_(*s*), 0 ≤ *s* < *t*}, the true time-to-conversion *T*^∗^ for patient *j,* and the sample (ADNI) on which the joint model was fitted:

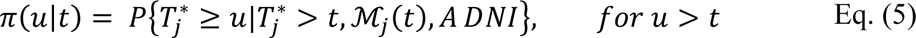

The probability in eq. (5) is named in the literature as a survival probability and can be estimated using methods described in (13), such as Empirical Bayes or Monte Carlo; *π*(. ) defines the risk of no-progression from MCI to AD and it is dynamic in the sense that, if the information is available at time t^′^ > *t*, *π*(. ) will be updated to *u* > *t*^′^.

#### (5) Assessment of prediction

It is well known that HC volumes decrease as ADAS-Cog scores increase in the context of AD. Consequently, the risk of conversion increases and AUC, the area under ROC curve also increases with larger HC atrophy.

The joint model can be used to provide cohort and individualized predictions for conversion to AD and HC as a longitudinal biomarker. To assess the quality of these predictions we present discrimination measures based on AUC.

If we have two subjects, one having and the second not having converted to AD, the AUC can be evaluated by the formula (26):

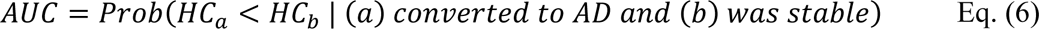

This postulates that for any two subjects (a,b), the AUC conveys the ranking of biomarker levels between subjects, concordant with their ranking for the conversion of MCI to AD. The formula in eq. (6) can also help us to select the best biomarker able to discriminate between progressive and stable subjects, given a biomarker of interest. An AUC = 1 indicates maximum discrimination, whereas AUC = 0.5 indicates random discrimination.

## 3. RESULTS

Socio-demographic and neuropsychological characteristics of the two groups of MCI subjects enrolled in the study are detailed in Table 1. At baseline, groups are significantly different in %F (*p < .0001*), %M (*p<.0001*), ADAS-Cog (*p<.0001*), but weakly in age (*p=0.0319*) and not in education p=0.541). Baseline ADAS-Cog scores ranged between 3 and 39.7 (inclusive) with a median of 16, while at last follow-up they were between 5 and 84 (inclusive) with a median of 32. At the time point of conversion, progressing MCI subjects had an ADAS-Cog score between 5 and 52 (inclusive) with a median of 26.3. The overall progression in ADAS-Cog therefore justified its use as a disease evolution proxy (time equivalent). The groups also differed (Table 1 & Figure 2) with respect to baseline bilateral HC volumes (*p* < .0001).

**Figure 2.**
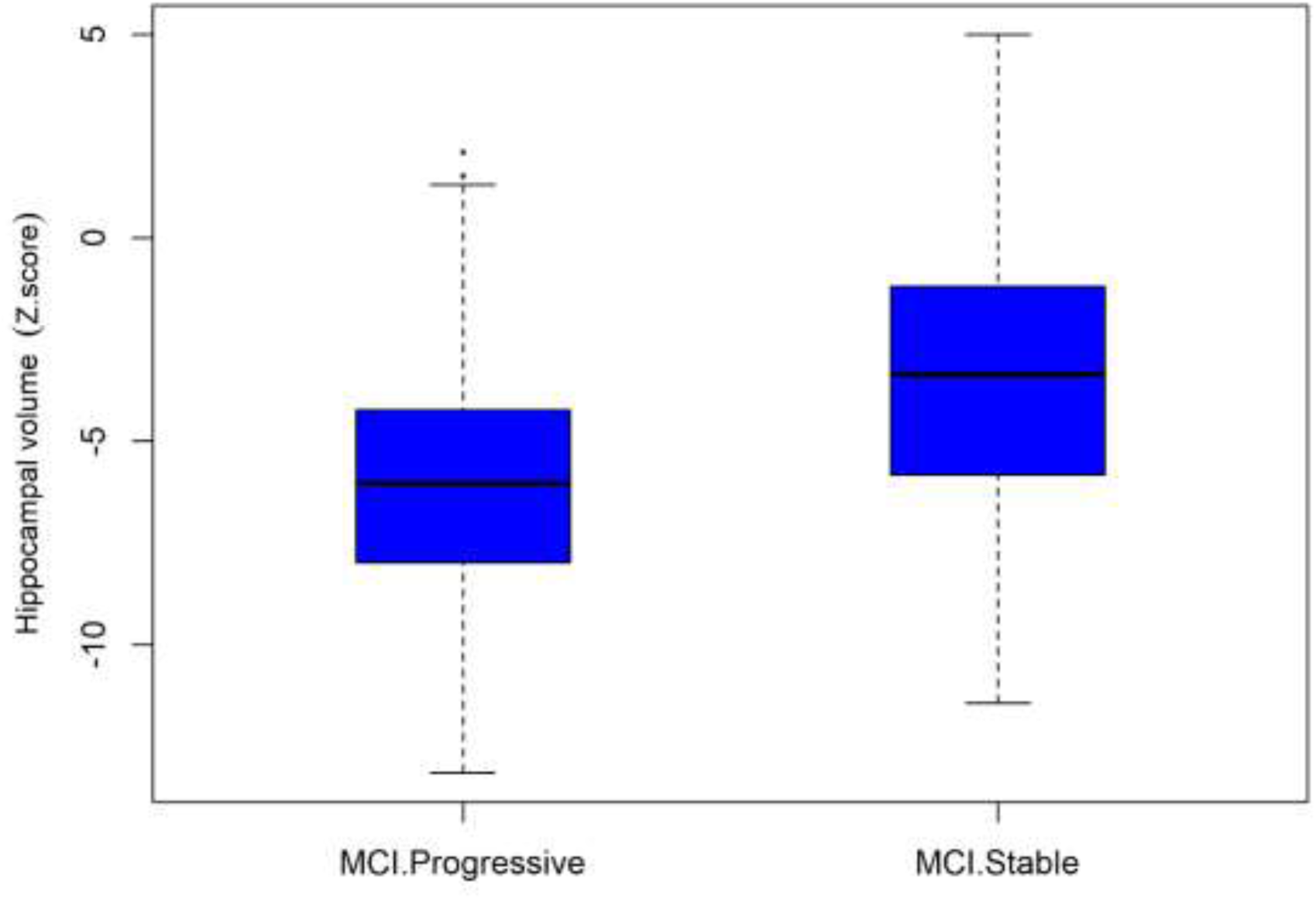
Progressive and stable MCI distributions for baseline adjusted hippocampal (HC) volumes.

**Table 1:** Subject group characteristics. p-value are obtained from t.test (or Wilcoxon test) and Chi-square tests, to compare Characteristics between MCIs and MCIp, depending on the nature and distribution of the data. First line represents the percent of MCIs and MCIp by sex over all data.

| Characteristics |  | Status=Stable MCI (n=529) | Status=Progressive MCI (n=285) | p-value |
| --- | --- | --- | --- | --- |
| Sex | Female (%) | 25.92% | 13.64% | <.0001 |
|  | Male (%) | 39.07% | 21.38% | <.0001 |
| Age (years) |  | 72.59 ± 7.83 | 73.74 ± 6.91 | 0.0319 |
| Education (years) |  | 15.99 ± 2.87 | 15.87 ± 2.76 | 0.541 |
| ADAS (score) |  | 14.27 ± 5.94 | 20.62 ± 6.26 | <.0001 |
| Bilateral HC (Z.score) |  | -3.49 ± 3.18 | -6.00 ± 2.83 | <.0001 |

**Table 2.**
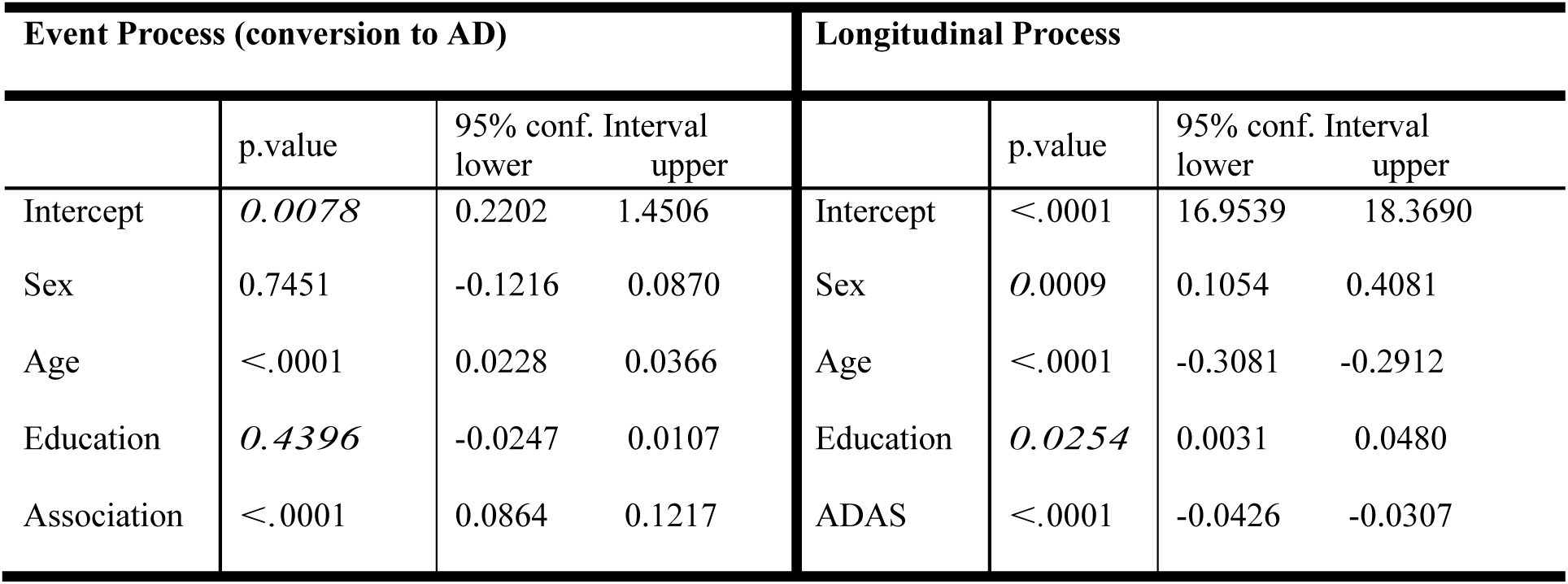
p-values under the joint modeling for event and longitudinal process.

In Figure 3, we present the Kaplan-Meier curve to estimate the global risk of conversion from MCI to probable AD. The curve shows that, as time increases (i.e. new follow-ups), the overall risk of conversion for the whole group increases; it further depends on HC volume change.

**Figure 3.**
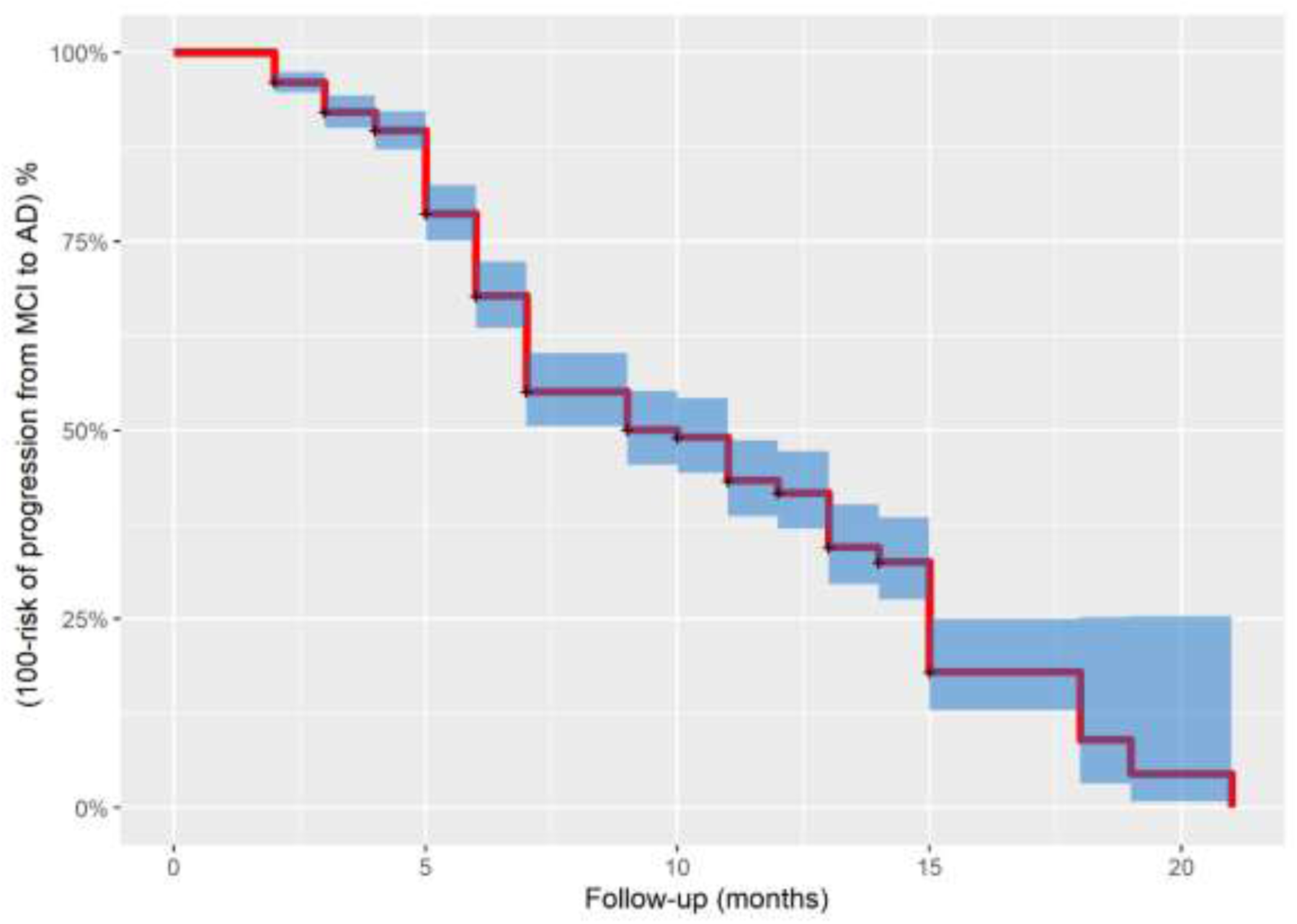
Kaplan Meier curves. As time increases, the average risk conversion increases for the whole group.

Our goal being to achieve a risk estimate for each subject, we wished to combine this global risk with the effect of longitudinal measures of bilateral HC volumes on the conversion process.

First, we had to ascertain if the two models were associated. The result of test the association parameter ***α*** (eq. 3) (see Table 2) was highly significant (*p* < .0001, C.I = [0.0864 ; 0.1217]), indicative of a strong association between HC volume changes and progression from MCI to AD. In the event process part of our joint model, Sex (*p=0.7451*) and education (*p=0.4396*) did not influence progression risk (see Table 2), however, age has a strong effect (p < .0001). The association *⍺* also indicates the strong effect (p < .0001) of HC evolution on the risk of conversion.

These postulates being verified, we therefore generated the joint model according to eq. 4. Figure 4 shows four randomly selected subjects from ADNI with their risk of conversion plotted against HC volume changes through time (as shown via the proxy surrogate marker). As can be expected, the risk of progression grows faster if HC volumes at baseline were lower than the mean, or if HC volumes decreased faster.

**Figure 4.**
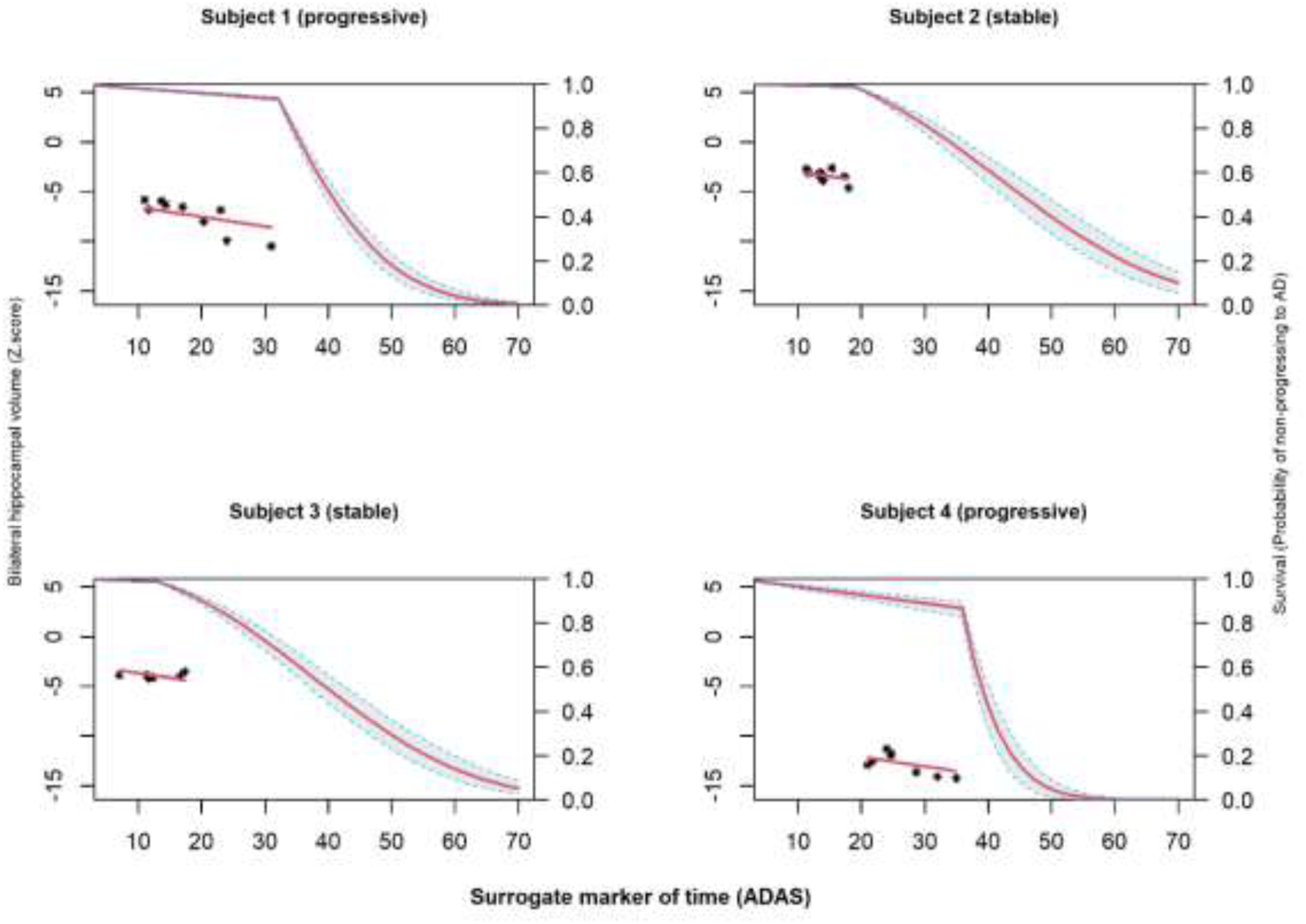
Conversion risk for four randomly selected subjects from ADNI. As HC volume decreases, the risk of conversion increases for each subject. Left side scale = HC volume Z-scores; right hand scale = risk of conversion. Stars indicate individual HC volume measurements. These four subjects were not included in the data used to fit joint model.

We can also estimate the risk of progression for new subjects at each timepoint, given new biomarker information, as illustrated in Figure 5.

**Figure 5.**
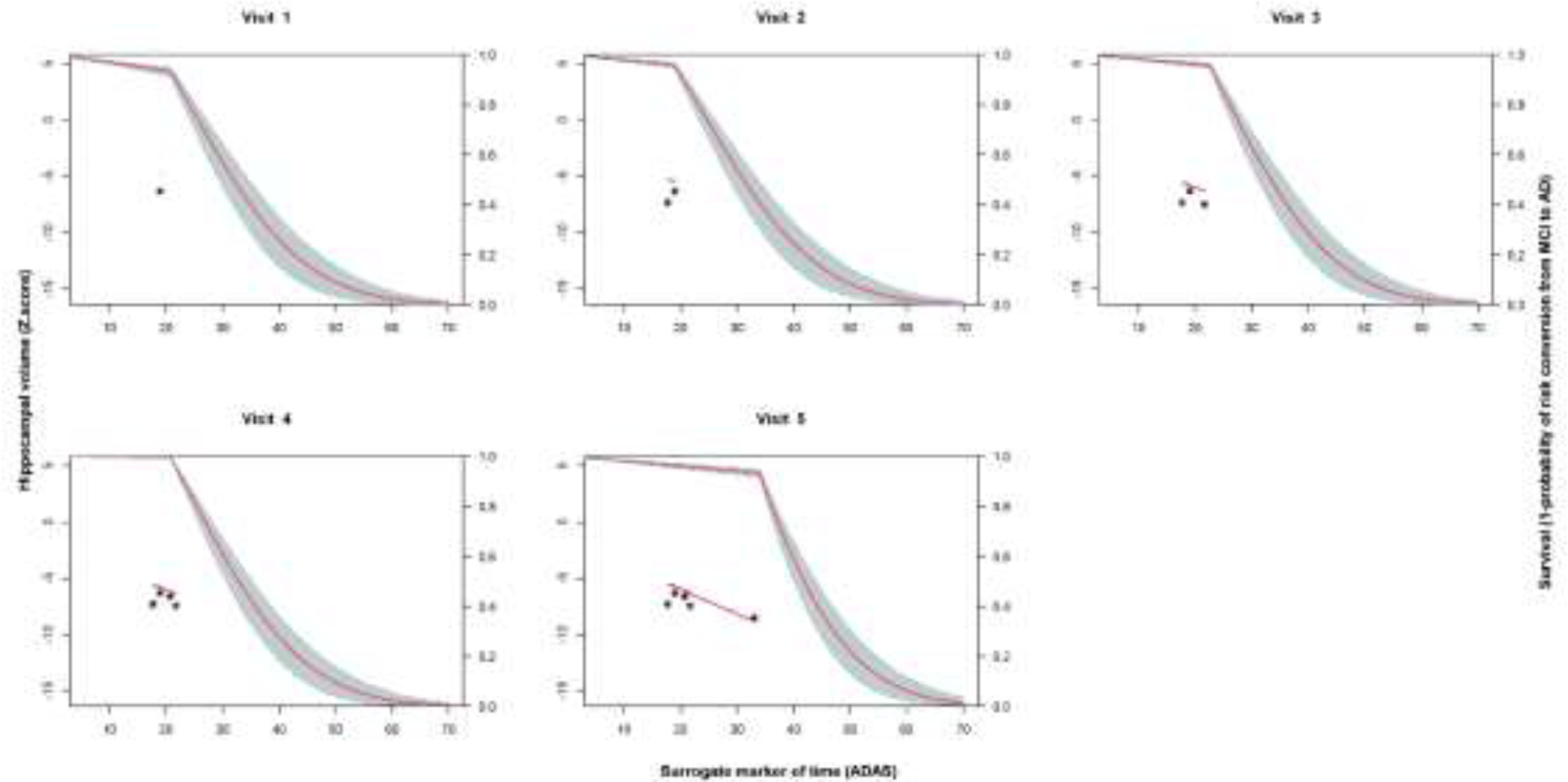
Dynamic prediction of risk conversion for a single subject who has six visits. As we have more information on the bilateral HC, the probability of risk conversion was updated. Left side scale = HC volume Z-scores; right hand scale = risk of conversion. Stars indicate individual HC volume measurements (up to six follow-ups in this example). This single subject was not included in the data used to fit joint model.

The accuracy of prediction is shown in Figure 6. At each follow-up, specifically after 6, 12, 24, or 48 months, the figure illustrates that as time increases, the AUC increases (Table 3). This indicates that the probabilities of the biomarker HC correctly classifying a subject as converted to AD or non-converted increase. The higher the AUC, the more accurate the predictions rule are.

**Figure 6.**
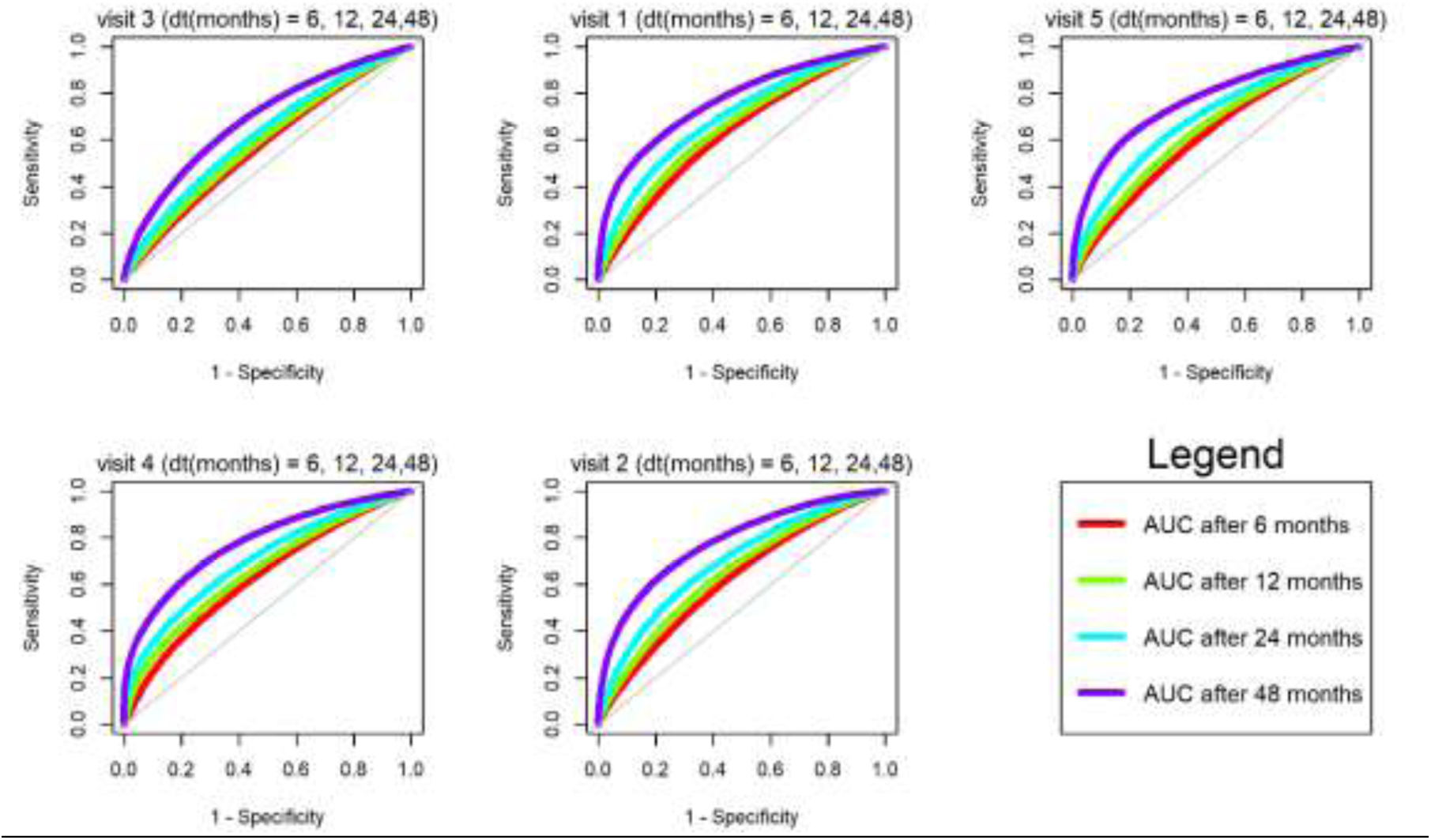
ROC curve and AUC for different variations of ADAS shows the accuracy of dynamic prediction. The values of different AUC are in. **table 3**.

**Table 3.** AUC over time after each visit.

| UC \ Time | Visit 1 | Visit 2 | Visit 3 | Visit 4 | Visit 5 |
| --- | --- | --- | --- | --- | --- |
| After 6 months | 0.587 | 0.636 | 0.627 | 0.637 | 0.629 |
| After 12 months | 0.603 | 0.663 | 0.654 | 0.669 | 0.658 |
| After 24 months | 0.633 | 0.704 | 0.700 | 0.712 | 0.704 |
| After 48 months | 0.702 | 0.774 | 0.779 | 0.788 | 0.789 |

The average conversion risk from MCI to AD for the complete stable and progressive groups are shown in Figure 7. One can appreciate that hippocampal volume decreases faster in the progression group, resulting in a higher risk compared to the stable group. This is confirmed by the higher association (see Table 2) between longitudinal and event process.

**Figure 7.**
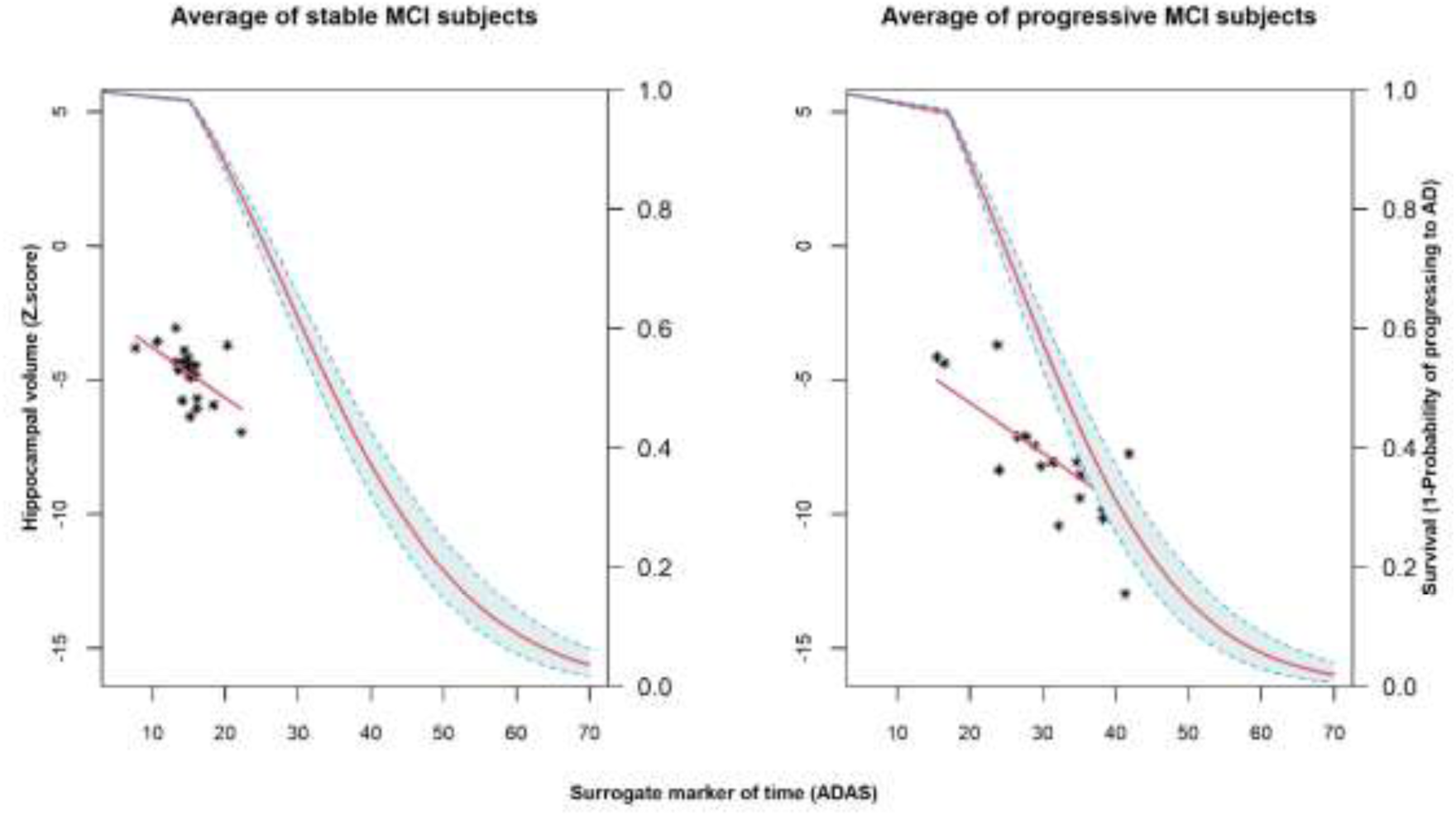
Prediction of global risk conversion for both groups of progressive and stable MCI subjects based on analysis of HC volumes. Left side scale = HC volume Z-scores; right hand scale = risk of conversion. Stars indicate individual HC volume measurements. Stars represent the mean of HC volumes in each group.

## 4. DISCUSSION

We have proposed a formulation for a longitudinal-survival joint model able to predict individual and global outcomes from biomarker data. Joint models correctly consider the relationship between the biomarker of interest (e.g., bilateral HC changes), and the fact that this biomarker is an internal (endogenous) covariate. In the ADNI dataset, we predicted conversion risk from MCI to AD using the evolution of hippocampal volume over time, with an AUC reaching 0.789 for the whole group, after 5 visits in 48 months.

### Joint approach for modelling

Joint models have been used in health studies (27) and offer advantages such as increasing the efficiency in statistical inference by utilising both the longitudinal and survival data simultaneously and understanding and quantifying the association between longitudinal and survival processes. By combining biomarker evolution and risk conversion, the resulting joint model bias is lower than in the classical approach (12), where either one is observed separately. Few have been applied to AD (e.g. (28)). Most are longitudinal models focusing on cortical thickness ((29), (30)), CSF ((31),(32)), FDG PET, ((33)), or in some instanced multiple factors (34). By conversion here is meant “cognitive trajectory leading to a diagnostic state”. The ADAS-Cog trajectory is useful clinically as once it reaches a certain level, it is indicative of objective impairment, then dementia. It will be obviously appreciated that our joint model approach is generalizable to other outcomes than conversion (e.g., response to therapy), as well as to other biomarkers within the context of AD (e.g., other cerebral imaging measures, CSF amyloid or tau), especially in this new era of amyloid-based disease-modifying therapies (35, 36), and can include different or more covariates beyond sex (e.g., APOE status) and/or other pathological outcomes. However, the extension to a multi-biomarker joint model is not as straightforward, as even univariate predictive models can become complex and carry a high computational cost.

### HC and ADAS-Cog as biomarkers of AD

Figure 7 shows that, on average, based on hippocampal volumes, progressive subjects (right side) decline faster than stable ones (left side). The risk of this conversion was estimated over the course of more than four-year follow-ups. The strong significant association between the evolution of hippocampal volume and the conversion risk shows that the latter risk depends on the trajectory of HC volume; hence, if hippocampal volumes decrease, the conversion risk increases both for the whole population and for one subject from this population.

Clinically, the relationship between MRI measures of temporal lobe structures, cognition and disease staging is strong, at least in the prodromal observable disease stages. It has been reported multiple times that MRI findings correlate with AD pathological progression. For example, Jack and colleagues (37) found an r^2^ = -0.63 for the association between bilateral hippocampal volume and Braak stages in AD patients, and r^2^ = 0.60 between bilateral hippocampal volume and Mini-Mental State Examination (MMSE) scores (similar in principle to ADAS-Cog); while Dawe et al. (38) observed a 0.2 z-score change in global cognition for each cm^3^ of hippocampal loss measured on MRI in a mixed study sample (cognitively normal controls, MCI and AD). Regarding the MMSE, Nelson and colleagues (39) reported correlations ranging from 0.58 to 0.82, according to age stratum, between the number of neurofibrillary tangles found at histopathology, and pre-mortem MMSE (40) scores. They further related that mean MMSE scores were between 25 and 28 in patients with Braak (41) I-III post-mortem AD changes (42). Hence, in a first approximation, using HC volume as a proxy of neurodegeneration due to AD and ADAS-Cog scores as a putative proxy for disease staging is of clinical relevance. Those markers bore the evident advantage in our case of having been collected, reliably, across the ADNI sample for follow-ups of up to 15 years, which is not the case for other markers (including markers of amyloid deposition); the major conclusion here being that joint survival-longitudinal models are demonstrably effective in this pathology, with these markers, and over this time frame. They can provide an individualized risk statistic at any given timepoint, which can be adjusted and refined as new information is provided.

### Comparison with other models

Other authors have compared joint models to separate analysis, such as an earlier article by Guo and colleagues, who proposed a joint model for longitudinal and survival data, obtaining maximum likelihood estimates via the EM algorithm, in the context of AIDS survival following HIV (e.g., (43)). We did not however find references for such models in the context of AD nor with our formulation.

## Conclusion

Individualized predictions of decline have great clinical importance. Statistical methods for survival models are well known and applicable however, may not be appropriate when the time-dependent covariate is internal and changes values between follow-up visits such as biomarkers. In this case, we propose joint statistical models that account for both longitudinal and time-to-event conversion simultaneously. In this article we developed the theoretical background and demonstrated its utility when applied to AD and based on HC volumes. This methodology can obviously be generalized to other outcomes and biomarkers.

## Data Availability

Disease Neuroimaging Initiative (ADNI) database (adni.loni.ucla.edu)

https://www.adni.loni.ucla.edu

## Abbreviations

AD: Alzheimer’s Disease
CTRL: Cognitively healthy control Subjects
HC: hippocampal volume
MCI: Mild Cognitive Impairment
ADAS-Cog: Alzheimer’s Disease Assessment Scale-Cognitive
MRI: Magnetic Resonance Imaging.

## AUTHOR CONTRIBUTIONS

A.M. conceptualized the study, analyzed the data, interpreted the results, and wrote the manuscript. O.P. revised the manuscript for important intellectual content. S.D. conceptualized the study, supervised the study process, and revised the manuscript for important intellectual content.

## STUDY FUNDING

We gratefully acknowledge financial support from the Alzheimer’s Society of Canada (#13-32), the Fonds de recherche du Québec – Santé / Pfizer Canada Discovery program, and the Canadian Institute for Health Research (#117121). S.D. is a Research Scholar from the Fonds de recherche du Québec – Santé (#30801).

## COMPETING FINANCIAL INTEREST STATEMENT

S.D. is officer and shareholder of True Positive Medical Devices inc. and was a paid consultant to Eisai.

## Notes

### Competing Interest Statement

The authors have declared no competing interest.

### Funding Statement

- Alzheimer's Society of Canada (#13-32);
- Fonds de recherche du Quebec - Sante / Pfizer Canada Discovery program, and the Canadian Institute for Health Research (#117121).

